# An Incognito Standardized Patient Approach for Measuring and Reducing Intersectional Healthcare Stigma

**DOI:** 10.1101/2023.08.21.23294305

**Authors:** M. Kumi Smith, Danyang Luo, Siyan Meng, Yunqing Fei, Wei Zhang, Joseph Tucker, Chongyi Wei, Weiming Tang, Ligang Yang, Benny L Joyner, Shujie Huang, Cheng Wang, Bin Yang, Sean Y Sylvia

## Abstract

**Background:** Consistent evidence highlights the role of stigma in impairing healthcare access in people living with HIV (PLWH), men who have sex with men (MSM), and people with both identities. We developed an incognito standardized patient (SP) approach to obtain observations of providers to inform a tailored, relevant, and culturally appropriate stigma reduction training. Our pilot cluster randomized control trial assessed the feasibility, acceptability, and preliminary effects of an intervention to reduce HIV stigma, anti-gay stigma, and intersectional stigma.

**Methods:** Design of the intervention was informed by the results of a baseline round of incognito visits in which SPs presented standardized cases to consenting doctors. The HIV status and sexual orientation of each case was randomly varied, and stigma was quantified as differences in care across scenarios. Care quality was measured in terms of diagnostic testing, diagnostic effort, and patient-centered care. Impact of the training, which consisted of didactic, experiential, and discussion-based modules, was assessed by analyzing results of a follow-up round of SP visits using linear fixed effects regression models.

**Results:** Feasibility and acceptability among the 55 provider participants was high. We had a 87.3% recruitment rate and 74.5% completion rate of planned visits (N=238) with no adverse events. Every participant found the training content “highly useful” or “useful.” Preliminary effects suggest that, relative to the referent case (HIV negative straight man), the intervention positively impacted testing for HIV negative MSM (0.05 percentage points [PP], 95% CI,-0.24, 0.33) and diagnostic effort in HIV positive MSM (0.23 standard deviation [SD] improvement, 95% CI, -0.92, 1.37). Patient-centered care only improved for HIV positive straight cases post-training relative to the referent group (SD, 0.57; 95% CI, -0.39, 1.53). All estimates lacked statistical precision, an expected outcome of a pilot RCT.

**Conclusions:** Our pilot RCT demonstrated high feasibility, acceptability, and several areas of impact for an intervention to reduce enacted healthcare stigma in a low-/middle-income country setting. The relatively lower impact of our intervention on care outcomes for PLWH suggests that future trainings should include more clinical content to boost provider confidence in the safe and respectful management of patients with HIV.

## INTRODUCTION

Stigma is one of the most commonly cited reasons for poor uptake of evidence-based interventions (EBI) for HIV prevention such as testing or preexposure prophylaxis.[1] Stigma impedes care access through several mechanisms. Patients who feel undeserving of care (internalized stigma) or who fear discrimination (anticipated stigma), for example, may abstain from seeking EBI, whereas clinical staff may discriminate by offering suboptimal care or turning patients away (enacted stigma). Uptake is lowest in key populations such as gay and bisexual men who have sex with men (MSM) who face additional stigma on account of social taboos against same-sex or other behaviors. The interaction of multiple stigmatized identities, or intersectional stigma, is an increasingly recognized aspect of HIV stigma, but few effective interventions exist.[2–6] [5,7] A better understanding of the layered nature of stigma is necessary for disentangling its effects and for informing design of effective interventions.[2,7,8]

Interventions on stigma, including those in healthcare settings, have been plentiful enough to motivate multiple reviews on the topic.[9–16] All the reviews note the abundance of impactful interventions, but caution that pervasive issues with design quality and methodological rigor limit meaningful insights as to what actually works. Central among the methodological issues is that of stigma measurement, itself a long-standing topic of discussion.[16–19] Enacted stigma in particular is singled out for its inherent challenges,[9,16] namely how few providers are likely to admit to discrimination and the difficulty of surveying patients who may lack the clinical knowledge to objectively evaluate their quality of care. Compounding these challenges is that of how to best measure intersectional stigma, as victims of this type of stigma cannot easily disentangle discrimination originating from different sources.[20]

Our team developed a stigma reduction training for providers by creating a novel approach to measuring enacted, intersectional stigma experienced by MSM, people living with HIV (PLWH), and people with both identities. We measure stigma by deploying incognito standardized patients (SP) or trained actors who present standardized disease cases for the purposes of clinical observation. Providers consent to visits in advance but are not told when they will take place, allowing insights into their true behaviors in real clinical settings. By randomly varying the sexual orientation and HIV status of presented cases, we can quantify stigma as the difference in care quality received across scenarios. Results are shared with separate community advisory boards (CAB) of local providers and MSM to solicit their views on stigma drivers, allowing us to enlist the insights of people closest to the problem.[21] We hypothesize that our intervention can more effectively reduce stigma by giving trainees more tailored and hands on content than traditional curricula informed by theoretical reasoning alone.[22–24] Given the importance of STI care as an entry point into the HIV prevention continuum for MSM at highest risk of HIV infection, we worked with providers of sexually transmitted infection (STI) clinics.

Results of the baseline round of stigma assessment using incognito SP visits are reported in a separate manuscript. Here we present the results of a pilot randomized control trial (RCT) to assess the feasibility, acceptability, and preliminary impact of a stigma reduction intervention informed by an unannounced SP approach to measuring stigma against MSM, PLWH, and people with both identities.

## METHODS

The pilot cluster RCT was conducted between March 2021 and August 2022. In brief, a round of unannounced SP visits were conducted at baseline, results of which were used to inform the design of the stigma reduction intervention. Following delivery of the training intervention, a second round of unannounced SP visits were conducted to assess the key outcomes of feasibility, acceptability, and preliminary impact of the intervention. An overview of the study and data collection procedures is provided in Figure 1.

**Figure 1.**
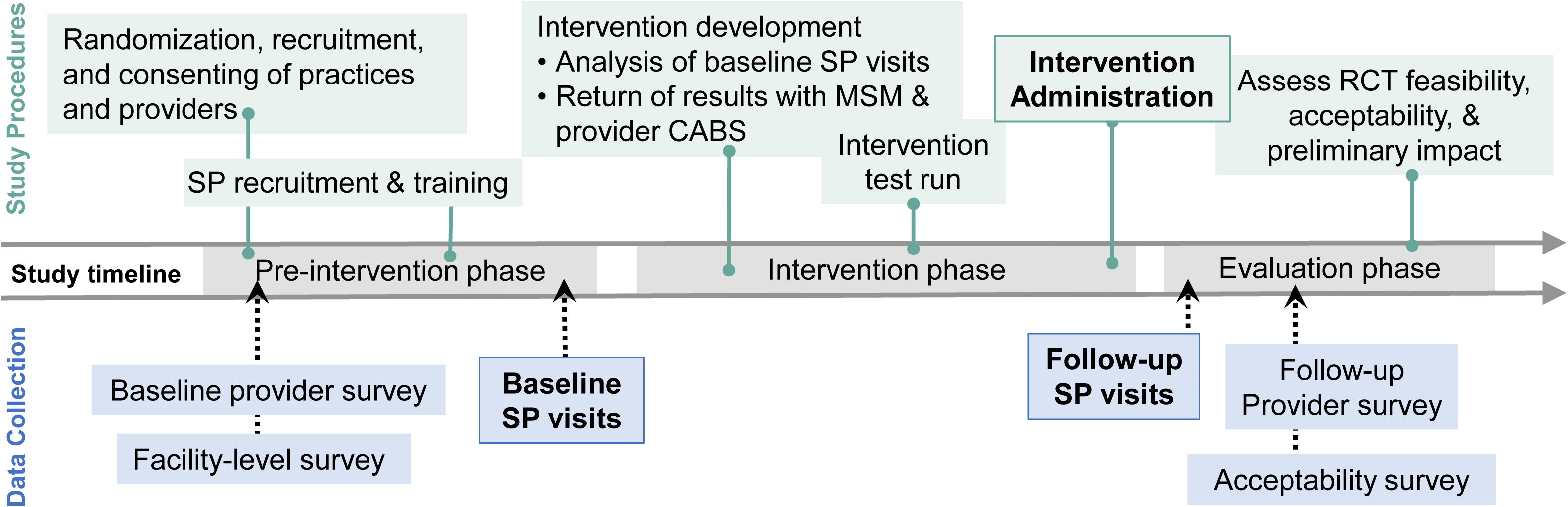
Study timeline, divided into procedures and data collection procedures.

### STUDY SETTING

This pilot RCT was conducted in Guangzhou, China. Guangzhou, a city of over 11 million residents, embodies the hallmarks of the Chinese HIV epidemic in MSM: rapidly rising HIV and STI prevalence and prevalent healthcare stigma.[25,26] The prefectural municipality of Guangzhou is made up of 11 urban districts, 10 from which we recruited our study clinics (Figure 2). All field activities were conducted in partnership with the Dermatology Hospital of the Southern Medical University (SMU) which oversees surveillance, clinical practice, and implementation of disease control policy through a province-wide network of >400 STI practices. Practices included both standalone clinics and specialty wards within larger hospitals.

**Figure 2.**
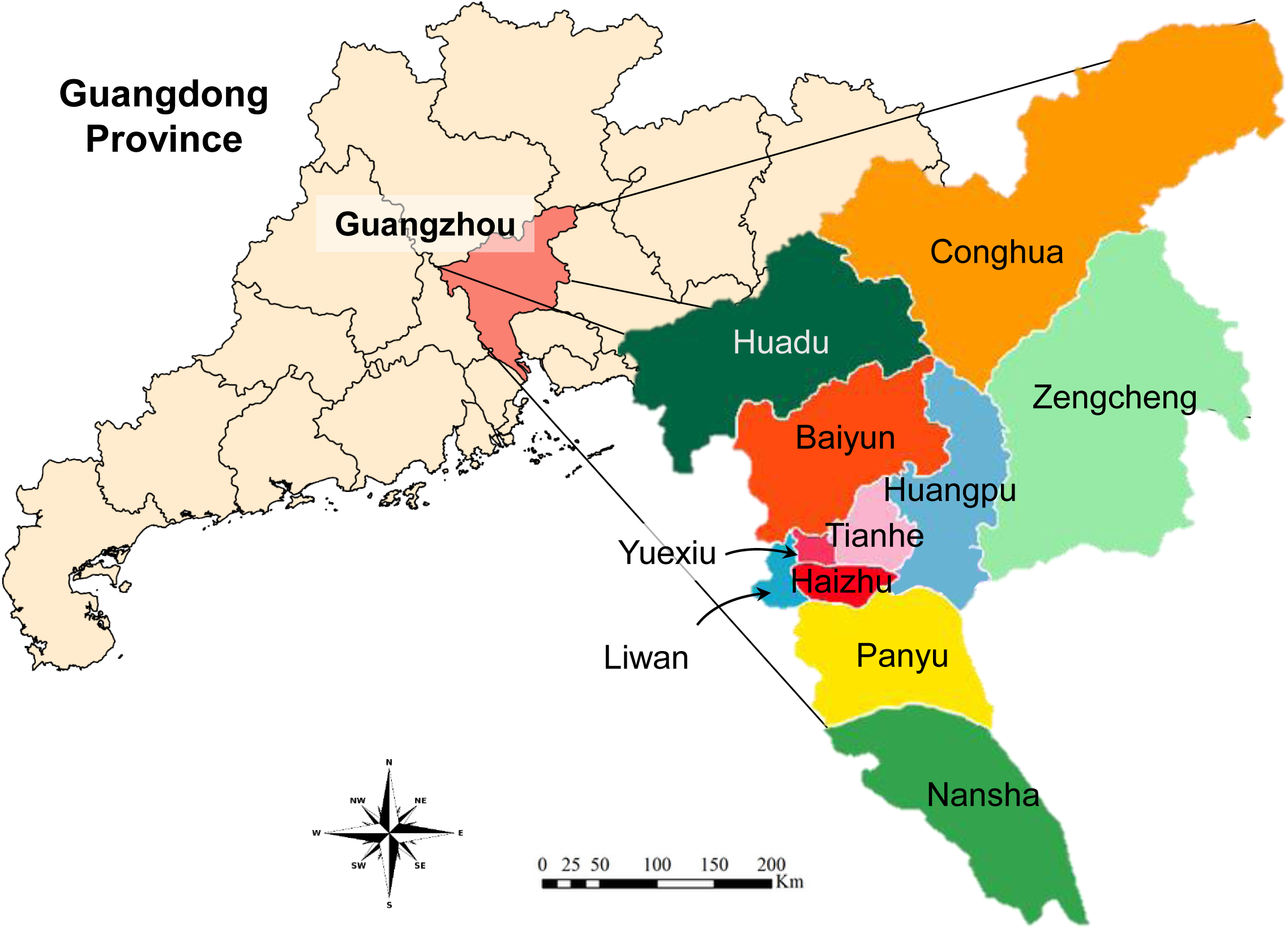
Map of the study setting. The prefectural municipality of Guangzhou is made up of 11 urban districts. Figure 3. CONSORT map of the pilot cluster randomized control trial to reduce enacted healthcare stigma.

**Figure 3.**
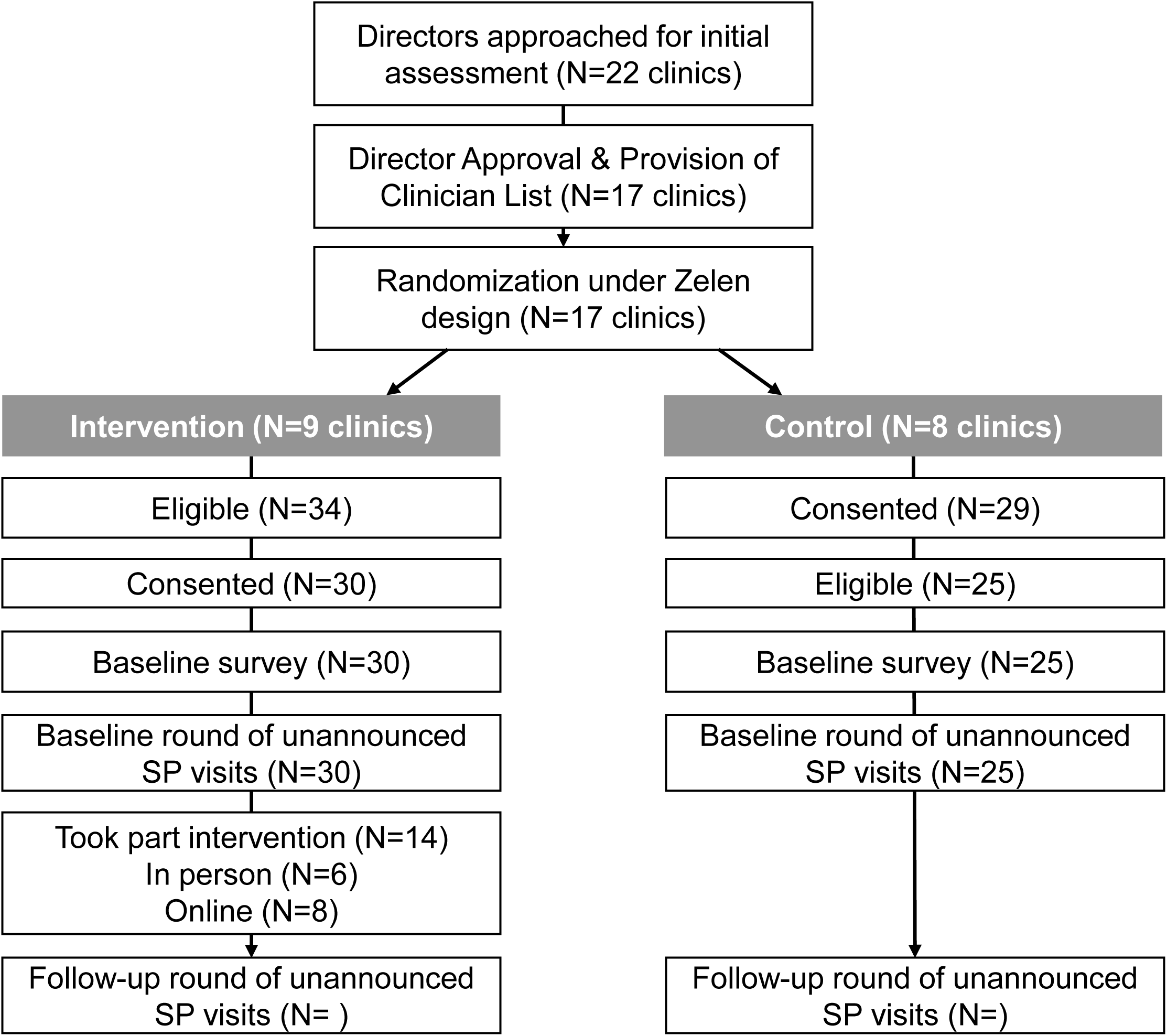
CONSORT map of the pilot cluster randomized control trial to reduce enacted healthcare stigma.

### RECRUITMENT & RANDOMIZATION

Our sampling frame consisted of STI practices listed on SMU’s network roster located in Guangzhou. The first stage of our two-step recruitment process consisted of approaching clinic or ward directors in person to explain the study goals and if they agreed, to obtain a list of providers employed at their practice. Eligible practices were those with formal government medical accreditation and with the capacity to provide enzyme-linked immunosorbent assay testing for HIV, treponemal (e.g., Treponema pallidum particle agglutination) and non-treponemal tests (e.g. rapid plasma reagin] for syphilis. In the second stage of recruitment we approached practice providers individually to inform them of study goals, answer questions, and obtain consent. Eligible providers were 1) at least 18 years of age; 2) certified to provide STI related care in Guangdong province, and 3) planning to remain at the practice clinic for at least one year.

Randomization at the practice level used a modified Zelen design in which control arm participants are not informed that they are part of an RCT.[27–29] This approach, which has been applied in fields ranging from STIs to chronic disease, seeks to minimize bias from potential compensatory behaviors of participants who are knowingly assigned to the control arm (i.e. the John Henry effect).[30] During consent procedures, all providers, regardless of arm, were instructed to document details of any suspected SP visits which could then be verified post study to assess the rate of SP detection by providers. All providers then took part in a 15-minute survey administered by trained study staff to provide information on their demographics and professional background. A facility level survey was also completed by appropriate clinic staff to document clinic characteristics such as staff size and patient load.

### DATA COLLECTION

We conducted two waves of incognito SP visits with consenting providers: one set at baseline and a second set four months after the intervention training. The presented case was a young male, age 20-40, presenting with complaints of primary syphilis (i.e. a recently healed chancre on the penis) and condomless sex. We chose primary syphilis because it allowed for plausible presentation by healthy volunteers and because of the public health significance of timely treatment for MSM and PLWH, both of whom experience elevated incidence relative to other populations. We randomly varied the HIV status and sexual orientation with by visit to obtain care quality measures on each of four scenarios: HIV negative straight man, HIV negative MSM, HIV positive straight man, and HIV positive MSM (scenarios are hereafter referred to as “referent,” “MSM only,” “HIV only”, and “intersectional). SPs announced the HIV status and sexual orientation at the top of each visit using scripted opening lines.

SP hiring and training were conducted in close collaboration with the Zhitong LGBT Center, a Guangzhou-based community-based organization (CBO) specializing in LGBT+ advocacy and health promotion. Candidate SPs who met the basic descriptions of the role took part in a two day training which sought to achieve 1) realistic and consistent case presentations across all SPs and 2) a consensus on interpretation of items on the healthcare quality checklist used for data collection. Training activities included a review of study materials (scripts, checklists, safety protocol), role plays, and field testing. A second fresher training was held prior to the follow-up round of visits.

Provider participants received three SP visits per wave. The following case features were randomized by facility and within each facility: the specific SP conducting the visit, the case scenarios presented, and the order in which each provider received the case scenarios. SP visits with the same provider were spaced out by a minimum of two weeks to reduce risk of SP detection. Immediately after each visit, accompanying study staff conducted data collection with SP using the healthcare quality checklist and a brief qualitative interview to capture visit features that might have been missed by the checklist. Throughout the study, SPs met periodically as a team to discuss checklists and ensure mutually consistent interpretations of items and ratings.

### MEASURES

Data collection was conducted using a healthcare quality checklist designed to capture multiple dimensions of provider behavior that could theoretically shape clinical and interpersonal patient experiences (see **Supplemental materials**). Clinical items were informed by national diagnosis and treatment guidelines on syphilis case management,[31] and interpersonal items by input from both of our CABs. Data collected using the healthcare quality checklist was then operationalized into stigma measures by estimating differences in the quality of care between each of the test cases and the referent case (straight, HIV negative). Care quality was measured across the three domains of care including syphilis testing, diagnostic effort, and patient-centered care, details of which are included in the **Supplemental materials.**

We also measured feasibility of the intervention as recruitment rates, retention rates, and incidence of adverse events. Acceptability was measured using responses to a self-administered online survey distributed to providers following final study visits. Respondents were asked to evaluate various components on a 4-point Likert scale ranging from “dissatisfied” to “very satisfied.”

### INTERVENTION DEVELOPMENT

Details of enacted healthcare stigma observed at baseline are reported in a separate manuscript currently under review. Briefly, baseline visits documented evidence of all three forms of stigma: HIV stigma, MSM stigma (or heterosexism), and intersectional stigma. Stigma was most apparent in the lower quality of clinical care (syphilis testing, diagnostic effort) received by the various scenarios, whereas patient-centered care scores were similar across all four scenarios. The study team, with input from CAB members, preliminarily concluded that stigma towards gay and HIV positive patients manifested in this setting as neglect or avoidance, most likely due to providers’ lack of knowledge or exposure to these types of patients.

Analysis of our baseline results and input from our two CABs were evaluated using the information, motivation, and behavioral skills (IMB) model of behavior change as a guiding framework.[32] The resulting intervention centered on the three following goals: 1) to convey the significance of the STI epidemic in marginalized populations including MSM and PLWH; 2) to persuade providers of the public health significance of their role; and 3) to strengthen their skills in communicating with marginalized patients. Components included didactic sessions supported by prepared animated videos, role plays with volunteer SPs uninvolved in study visits, and group discussions on strategies to improve patient-provider communication. To accommodate provider schedules, a fully online version of the training was offered to providers who could not attend the in-person event.

#### Analysis

Preliminary effects of the pilot intervention were calculated using an intent-to-treat (ITT) approach. Our primary outcomes of MSM stigma, HIV stigma, and intersectional stigma were conceptualized as the difference in care quality received in each test scenario (i.e. MSM only, HIV only, intersectional) and the referent scenario (i.e. HIV negative straight man). Preliminary intervention effects were calculated for each domain of care: syphilis testing, diagnostic effort, patient-centered care. Linear ordinary least squares models were used to estimate training effects for each of the primary outcomes as follows:

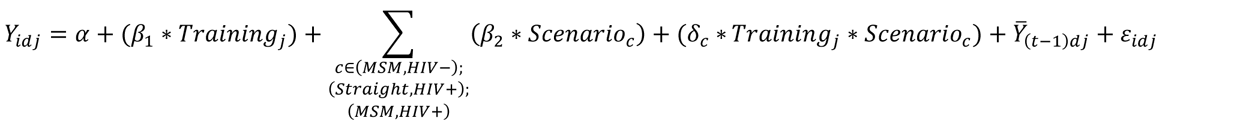

where *Y*_*idj*_ is a given post-intervention outcome for a clinical encounter between an SP with doctor *d* in facility *j*; *training* is a binary indicator for study arm assignment (1 for facilities randomized to the training intervention and 0 for control facilities); and *Scenarioc* is a set of binary indicators to designate the presented test scenario. To enhance statistical precision, each model controlled for the average of the outcome in all interactions with doctor *d* at baseline, ^*Ȳ*^_(*t*−1)*dj*_. As a cluster RCT, randomization was conducted by facility; thus our heteroskedasticity-consistent standard errors adjusted for clustering at the facility level.[33,34]

This regression specification yields estimates for *δ*_*c*_, three primary quantities of interest for each outcome. As increases in each outcome represent better care, this measures the extent to which differences in care relative to the referent group changed due to the intervention. Estimates above the null value of 0 represent a reduction in enacted stigma. Estimates below the null correspond to a smaller improvement in care quality relative to the referent scenario and are interpreted as *increase*s in a particular form of stigma.

#### Ethical Approvals

This study was approved by the institutional review boards at the University of Minnesota, University of North Carolina, and the Dermatology Hospital of the Southern Medical University. All study participants provided written informed consent in Chinese. Our study followed the Consolidated Standards of Reporting Trials (CONSORT) reporting guideline.

## RESULTS

### Sample Characteristics & Follow-up

The CONSORT flow diagram (Figure 3)[35] shows the recruitment and retention patterns for the study. According to the Zelen design, randomization occurred before providers were approached for individual consent. Following randomization, all eligible providers in each clinic were approached by study staff. Of the 34 who were approached at intervention arm clinics, 30 agreed to participate (88.2%); and of the 29 in the control arm, 25 (86.2%) agreed.

Providers had a mean age of 42 (standard deviation [SD], 9), were mostly male (62%), and were mostly assistant-or intermediate level clinicians (63%, as opposed to associate-or senior-level clinicians; Table 1). Enrolled clinics had a mean patient load of 861 weekly outpatients (SD 579) and employed an average of 4.8 clinicians (SD 2.9) and 2.2 (SD 2.6) support staff (Table 2).

**Table 1.** Provider characteristics participating in the study in Guangzhou, China.

|  | Total | Control | Treatment | Difference<sup>1</sup> | 95% CI<sup>1</sup> | p-value<sup>1</sup> |
| --- | --- | --- | --- | --- | --- | --- |
| Total (N) | 55 | 25 | 30 |  |  |  |
| Provider Age, Mean (SD) | 42 (9) | 43 (9) | 40 (9) | 2.9 | -1.9, 7.8 | 0.2 |
| Gender, n (%) |  |  |  | 0.07 | -0.46, 0.60 |  |
| Female | 21 (38.2) | 10 (40) | 11 (36.7) |  |  |  |
| Male | 34 (61.8) | 15 (60) | 19 (63.3) |  |  |  |
| Education, n (%) |  |  |  | 0.43 | -0.11, 0.97 |  |
| Professional School | 2 (3.6) | 0 (0) | 2 (6.7) |  |  |  |
| Bachelor Degree | 13 (23.6) | 5 (20) | 8 (26.7) |  |  |  |
| Graduate Degree | 40 (72.7) | 20 (80) | 20 (66.7) |  |  |  |
| Title, n (%) |  |  |  | 0.65 | 0.11, 1.2 |  |
| Assistant-level clinician | 10 (18.2) | 3 (12) | 7 (23.3) |  |  |  |
| Intermediate-level clinician | 25 (45.4) | 12 (48) | 13 (43.3) |  |  |  |
| Associate-level clinician | 16 (29) | 7 (28) | 9 (30) |  |  |  |
| Senior clinician | 3 (5.5) | 3 (12) | 0 (0) |  |  |  |
| Other | 1 (1.8) | 0 (0) | 1 (3.3) |  |  |  |
| Average Work Hours per Week, Mean (SD) | 39.6 (5.4) | 39.0 (4.0) | 40.0 (6.4) | -0.96 | -3.8, 1.9 | 0.5 |
| Average Patient Load per Week, Mean (SD) | 49 (20) | 41 (18) | 55 (20) | -14 | -24, -3.7 | 0.009 |

<sup>1</sup> Welch Two Sample t-test; Standardized Mean Difference
CI: Confidence Interval

**Table 2.** Clinic characteristics.

|  | **Total** | **Control** | **Treatment** | **Difference<sup>1</sup>** | **95% CI<sup>1</sup>** | **p-value<sup>1</sup>** |
| --- | --- | --- | --- | --- | --- | --- |
| Outpatients in past week, Mean (SD) | 861 (579) | 766 (614) | 944 (570) | -178 | -795, 439 | 0.5 |
| Number of clinicians, Mean (SD) | 4.9 (2.9) | 4.88 (3.7) | 4.9 (2.2) | -0.01 | -3.3, 3.3 | >0.9 |
| Number of clinicians above associate level, Mean (SD) | 2.1 (2.1) | 2.25 (2.6) | 2.0 (1.7) | 0.25 | -2.1, 2.6 | 0.8 |
| Number of support staff, Mean (SD) | 2.2 (2.6) | 1.63 (1.8) | 2.7 (3.1) | -1 | -3.7, 1.6 | 0.4 |
| Clinic provides treatment for occupational HIV exposure, n (%) | 16 (94%) | 8 (100%) | 8 (89%) | 11% |  | 0.6 |
| Clinic provides clinician training on patient-centered care, n (%) | 14 (88%) | 6 (75%) | 8 (100%) | -25% |  | 0.3 |
| Unknown | 1 | 0 | 1 |  |  |  |

1. Welch Two Sample t-test; 3-sample test for equality of proportions without continuity correction
CI: Confidence Interval

A baseline round of 123 unannounced SP visits were successfully completed: 72 in the intervention arm, 51 in the control arm. Of the 165 total planned visits, 123 (74.5%) were completed, for an 80% completion rate in the intervention arm and 68% in the control arm. Four months following intervention completion, we completed 115 (69.9%) of the 165 planned second wave visits: 71.1% in the intervention arm and 68% in the control arm. Reasons for non-completion included providers being on temporary leave (e.g. medical, maternity), leaving their position at the clinic, or being unavailable for visits after two attempts. No adverse events were reported during any of the visits. 41 (87.2%) of providers did not suspect or did not know if they had received an SP visit. None of the remaining 6 were able to recall visit dates, precluding our ability to verify SP detection.

### Feasibility & Acceptability

Regarding feasibility, 77.3% of the 22 clinic directors we approached agreed to participate in our study (N=17), and 87.3% of the 66 individual providers approached enrolled in the study (N=55). 14 of the 30 participants in the treatment arm took part in the intervention (46.7%), 6 in the in-person training and 8 in the synchronous online intervention. The remainder (41.7%) received intervention materials via Wechat (a popular text messaging app). The most commonly reported reasons for not attending the intervention included time conflicts, not having enough time, and facing unexpected COVID related travel restrictions.

Regarding acceptability of the intervention, all who took part in either in-person or online training reported that each training component (didactic, role play, group discussion) was “very useful” or “quite useful” (as opposed to “a little bit useful” and “not useful”). The portion that reported content as “very useful” was higher for the in-person attendees (66.7-83.3%) than for online attendees (37.5%). Similar patterns were observed in reported rates of satisfaction with aspects of the training delivery including pacing, difficulty, and quality of material, as well as the knowledge and preparation of trainers.

### Preliminary effect

Estimates of the marginal intervention effects on each type of stigma with each of the three domains of care are shown in Figure 4. In terms of syphilis testing, the intervention had a modest positive impact on MSM stigma (0.05 percentage points [PP]; 95% confidence interval [CI],-0.24, 0.33) and negatively impacted HIV and intersectional stigmas (-0.23 PP, 95% CI,-0.55, 0.085 and -0.07 PP; 95% CI, -0.38, 0.25, respectively), though all of these estimates lacked statistical precision. In terms of diagnostic effort, the intervention had negative impacts on MSM and HIV stigma (SD, -0.45, 95% CI, −1.60, 0.69; SD, −0.97, 95% CI, -1.83, -0.11, respectively) and a positive impact on intersectional stigma (SD, 0.23; 95% CI, -0.92, 1.37), though only the estimate for HIV stigma was statistically significant. Lastly for patient-centered care, we observed near null effects for MSM and intersectional stigma (SD, -0.01, 95% CI, - 0.83, 0.82; SD, 0.00; 95% CI, -1.15, 1.14) and a positive impact on HIV stigma (SD, 0.57; 95% CI, - 0.39, 1.53), though once more all estimates lacked statistical precision.

**Figure 4.**
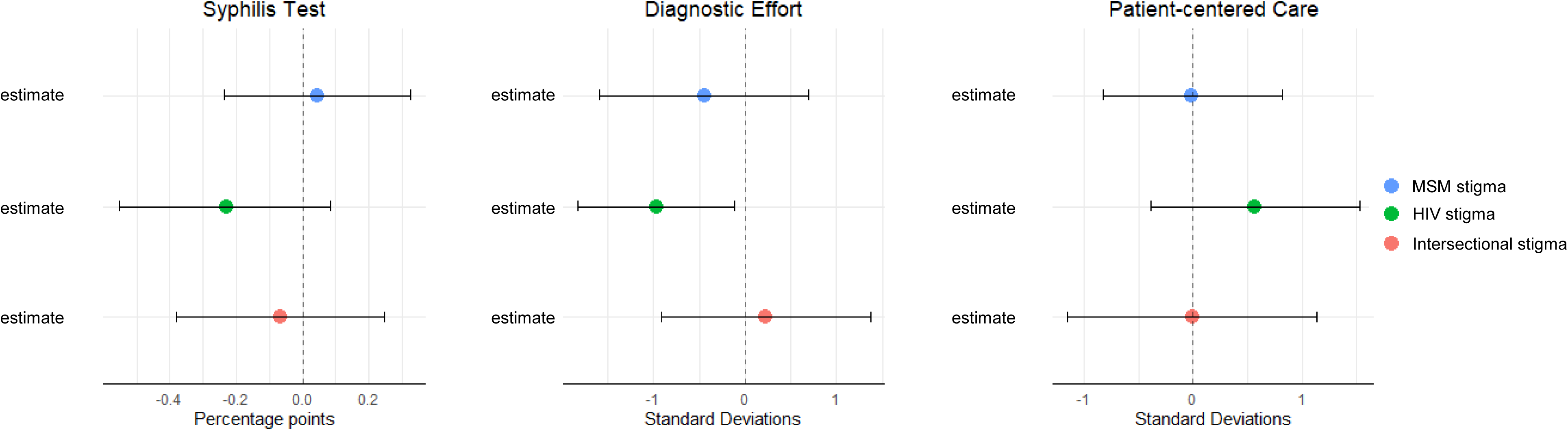
Marginal treatment effects for each of the three primary outcomes. Estimates reflect changes in pre-post stigma measures for each scenario (i.e. HIV negative MSM; HIV positive straight man; HIV positive MSM) relative to the same changes in the referent scenario (HIV negative straight man).

Additional insights are provided by stigma-specific estimates which quantify the absolute (vs. relative) impact of the intervention impact on each domain of care (Figure 5). These results indicate that the intervention had an absolute positive impact on the probability of syphilis testing for HIV negative MSM, the amount of diagnostic effort invested in HIV positive MSM, and the patient-centeredness of care for HIV positive MSM, though all of these estimates lacked statistical precision.

**Figure 5.**
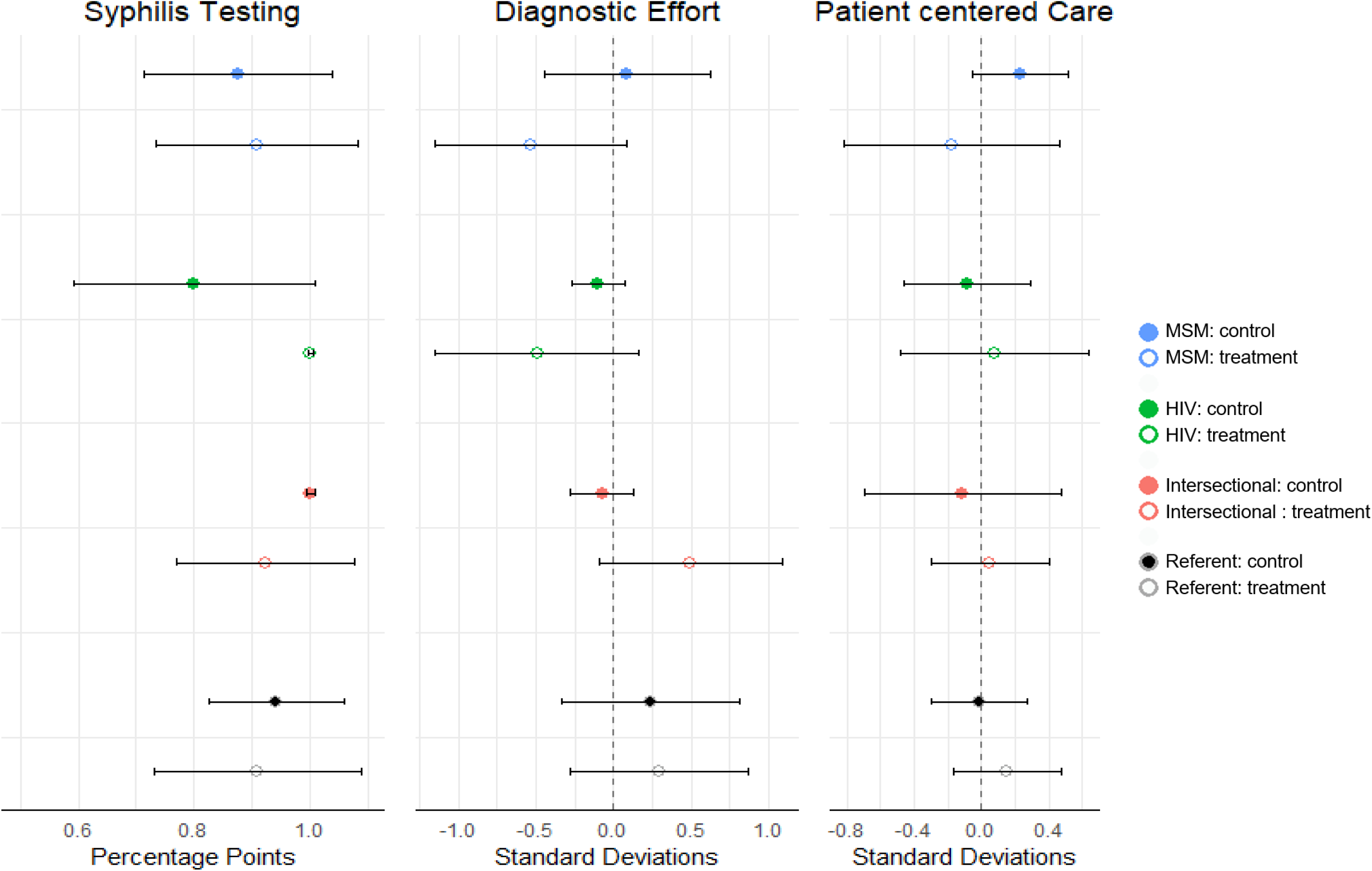
Scenario-specific marginal treatment effects.

## DISCUSSION

This pilot RCT documented high feasibility, acceptability, and several areas of impact for an intervention to reduce enacted healthcare stigma in a low/middle-income setting. The incognito SP approach provided unique insights into the particular ways that HIV stigma, MSM stigma, and intersectional stigma manifest in clinical settings, facilitating the creation of an intervention more responsive to providers’ actual service gaps and training needs. The objectivity of the incognito SP measure also allowed for a more rigorous evaluation of program impact. Our findings build off the one other known application of the incognito SP approach to measure enacted healthcare stigma, in which Li et al. dispatched SPs to compare behaviors of providers assigned to treatment versus control arms of an HIV stigma training in China.[36]

A central feature of our intervention was its distinct impacts on cases of different sexual orientations. That is, it appeared to improve clinical care—i.e. syphilis testing, diagnostic effort—for MSM of either HIV status but not for straight PLWH. This may be partially due to the mixing of our intervention message with those of our collaborators at SMU who as provincial STI authorities regularly emphasize the importance of MSM-facing clinical care to combat the regional syphilis epidemic. In addition, the siloed nature of China’s STI and HIV care systems may mean that our providers are far less likely to encounter patients who are PLWH than MSM. Due to their low exposure to PLWH—and lack of HIV related training—non-HIV specialists may therefore resort to avoidance or needless referral in their rare encounters with an HIV patient.[37–39] However the fact that treatment arm providers’ patient-centered care scores improved for HIV positive scenarios is an encouraging sign of their general receptiveness for ways to improve care for PLWH.

Though preliminary, results of this pilot RCT provide valuable guidance for future interventions and stigma research. First, the siloed nature of care for PLWH, an initially useful strategy to rapidly roll-out HIV treatment in many LMIC, may have also inadvertently deprived non-specialists of much needed training and experience in managing PLWH. This worsens stigma when non-specialists who feel underprepared resort to the understandable but problematic habit of perfunctory visits or needless referral. Future trainings should therefore feature relevant clinical skills in the correct, safe, and respectful management of patients with HIV. Second, our study exposed the challenges of targeting multiple forms of stigma, particularly when one identity (same sex behaviors among men) is a clinically valid risk factor for the other (HIV infection). Though some amount of trait-based generalization has a role in good clinical practice[40] excessive profiling can strain patient-provider relations.[41–43] Future trainings must therefore navigate the balance between healthy and harmful use of clinically salient patient history to improve care quality for key populations.

Findings from this pilot RCT should be interpreted in light of several key limitations. First, though our study was not powered to detect intervention effects, statistical power could have been strengthened by better participation in the intervention. Suboptimal participating was due in part to COVID-19 related prevention and reporting duties which consumed much of the limited free time our participants had. Many were restricted from travel due to COVID-related lockdowns. Future interventions may address these issues by dispatching academic details to deliver intervention content at each clinic. Second, our measure of patient-centered care relied on subjective assessments by individual SPs. Our SP training included team exercises to align their interpretation of items and rating scales across team members, but future uses of the unannounced SP approach may benefit from additional booster trainings to improve inter-reliability of SP reporting in order to improve data validity.

This study provides valuable proof of concept for the safe use of incognito SP visits for assessing, developing, and evaluating effective interventions to reduce enacted healthcare stigma in Chinese STI settings. The incognito SP approach is particularly well suited to measuring the more subtle and indirect forms of enacted stigma which may be less perceptible to individual patients, rendering patient surveys less reliable. The approach is also highly adaptable for capturing stigma from other sources (e.g. race/ethnicity, gender, sex, age) and a potentially powerful to assess the impact of structural stigma[44,45] by including facility-level features (e.g. support staff attitudes, clinic practices) as part of SP data collection. Finally, SP approaches create opportunities for meaningful co-creation of interventions with both stigmatized communities (MSM) and the intervention targets (providers), furthering the principles of community based participatory research.[46]

## AUTHOR CONTRIBUTIONS

M.K.S. and S.Y.S. conceived the study. M.K.S., S.Y.S., D.L, S.M, J.T, W.T, L.Y, B.L.J, S.H, C.W and

B,Y collaborated in the design of the methods and intervention. M.K.S. and S.Y.S. conducted the analyses with help from D.L and S.M. M.K.S wrote the initial drafts. S.Y.S provided inputs for the revision, J.T, W.T provide comments for the draft. All authors read and approved the final manuscript.

## Data Availability

All data produced in the present study are available upon reasonable request to the authors

## ACKNOWLEDGEMENTS

We thank all hospitals and providers for taking part in this study, as well as the The Guangdong STD Control Center. We also thank the Zhitong LGBT center for their support and connection to their community members. We would also like to express our gratitude for the hard work of all the SPs.

## FUNDING

This research was supported by NIH grants R34MH121251 (MKS and SYS)

**Trials Registration**: NCT04896216

## REFERENCES

1. Sullivan MC, Rosen AO, Allen A, Benbella D, Camacho G, Cortopassi AC, et al. Falling Short of the First 90: HIV Stigma and HIV Testing Research in the 90–90–90 Era. AIDS Behav. 2020 Feb 1;24(2):357–62.

2. Friedland BA, Sprague L, Nyblade LC, Baral SD, Pulerwitz J, Gottert A, et al. Measuring intersecting stigma among key populations living with HIV: implementing the people living with HIV Stigma Index 2.0. JAIDS J Acquir Immune Defic Syndr. 2018;21(Supp 5):e25131.

3. Rausch D. Promoting Reductions in Intersectional StigMa (PRISM) to Improve the HIV Prevention Continuum. 2018.

4. Rice WS, Logie CH, Napoles TM, Walcott M, Batchelder AW, Kempf MC, et al. Perceptions of intersectional stigma among diverse women living with HIV in the United States. Soc Sci Med. 2018;

5. Logie CH, James Ll, Tharao W, Loutfy MR. HIV, gender, race, sexual orientation, and sex work: A qualitative study of intersectional stigma experienced by HIV-positive women in Ontario, Canada. PLoS Med. 2011;8(11).

6. Goodin BR, Owens MA, White DM, Strath LJ, Gonzalez C, Rainey RL, et al. Intersectional health-related stigma in persons living with HIV and chronic pain: implications for depressive symptoms. AIDS Care - Psychol Socio-Med Asp AIDSHIV. 2018;

7. Nyblade LC. Measuring HIV stigma: Existing knowledge and gaps. Psychol Health Med. 2006;11(3):335–45.

8. Reidpath DD, Chan KY. A method for the quantitative analysis of the layering of HIV-related stigma. AIDS Care. 2005;17(4):425–32.

9. Smith MK, Xu RH, Hunt SL, Wei C, Tucker JD, Tang W, et al. Combating HIV stigma in low-and middle-income healthcare settings: a scoping review. J Int AIDS Soc. 2020;23(8):e25553.

10. Nyblade L, Stangl A, Weiss E, Ashburn K. Combating HIV stigma in health care settings: What works? J Int AIDS Soc. 2009;12(1).

11. Sengupta S, Banks B, Jonas D, Miles MS, Smith GC. HIV interventions to reduce HIV/AIDS stigma: A systematic review. AIDS Behav. 2011;15(6):1075–87.

12. Feyissa GT, Lockwood C, Woldie M, Munn Z. Reducing HIV-related stigma and discrimination in healthcare settings: A systematic review of quantitative evidence. PLOS ONE. 2019 Jan 25;14(1):e0211298.

13. Brown L, Macintyre K, Trujillo L. Interventions to reduce HIV/AIDS stigma: what have we learned? AIDS Educ Prev. 2003/03/12 ed. 2003;15(1):49–69.

14. Mak WWS, Mo PKH, Ma GYK, Lam MYY. Meta-analysis and systematic review of studies on the effectiveness of HIV stigma reduction programs. Soc Sci Med. 2017;188:30–40.

15. Stangl AL, Lloyd JK, Brady LM, Holland CE, Baral SD. A systematic review of interventions to reduce HIV-related stigma and discrimination from 2002 to 2013: how far have we come? J Int AIDS Soc. 2013;16(3 Suppl 2).

16. Nyblade L, MacQuarrie K. Can We Measure HIV/AIDS-Related Stigma and Discrimination? Current Knowledge about Quantifying Stigma in Developing Countries. Usaid. 2006;(January):1– 21.

17. Earnshaw VA, Chaudoir SR. From conceptualizing to measuring HIV stigma: A review of HIV stigma mechanism measures. AIDS Behav. 2009;13(6):1160–77.

18. Karver TS, Atkins K, Fonner VA, Rodriguez-Diaz CE, Sweat MD, Taggart T, et al. HIV-Related Intersectional Stigma and Discrimination Measurement: State of the Science. Am J Public Health. 2022;112(S4):S420–32.

19. Mahendra VS, Gilborn L, Bharat S, Mudoi RJ, Gupta I, George B, et al. Understanding and measuring AIDS-related stigma in health care settings: A developing country perspective. Sahara J. 2007 Aug;4(2):616–25.

20. Harnois CE, Bastos JL. The promise and pitfalls of intersectional scale development. Soc Sci Med. 2019 Feb;223:73–6.

21. Li L, Wu Z, Liang LJ, Lin C, Guan J, Jia M, et al. Reducing HIV-related stigma in health care settings: a randomized controlled trial in China. Am J Public Health. 2013;103(2):286–92.

22. Sengupta S, Banks B, Jonas D, Miles MS, Smith GC. HIV interventions to reduce HIV/AIDS stigma: A systematic review. AIDS Behav. 2011;15(6):1075–87.

23. Mahajan APA, Sayles JN, Patel V a, Remien RH, Szekeres G, Coates TJ. Sigma in the HIV/AIDS epidemic: A review of the literature and recommendations for the way forward. Aids. 2008;22(Suppl 2):S67–79.

24. Nyblade L, Stangl A, Weiss E, Ashburn K. Combating HIV stigma in health care settings: What works? J Int AIDS Soc. 2009;12(1).

25. Cao B, Zhao P, Bien C, Pan S, Tang W, Watson J, et al. Linking young men who have sex with men (YMSM) to STI physicians: A nationwide cross-sectional survey in China. BMC Infect Dis. 2018;18(1):228.

26. Tang W, Mao J, Tang S, Liu C, Mollan K, Cao B, et al. Disclosure of sexual orientation to health professionals in China: Results from an online cross-sectional study. J Int AIDS Soc. 2017;20(1):1– 9.

27. Zelen M. A New Design for Randomized Clinical Trials. N Engl J Med. 1979 May;300(22):1242–5.

28. Homer CSE, E HCS. Using the Zelen design in randomized controlled trials: debates and controversies. J Adv Nurs. 2002 Apr;38(2):200–7.

29. Torgerson DJ, Roland M. What is Zelen’s design? BMJ. 1998 Feb;316(7131):606–606.

30. Schellings R, Kessels AG, ter Riet G, Knottnerus JA, Sturmans F. Randomized consent designs in randomized controlled trials: Systematic literature search. Contemp Clin Trials. 2006;27(4):320–32.

31. Chen ZQ, Zhang GC, Gong XD, Lin C, Gao X, Liang GJ, et al. Syphilis in China: results of a national surveillance programme. The Lancet. 2007 Jan;369(9556):132–8.

32. Fisher JD, Fisher WA, Williams SS, Malloy TE. Empirical tests of an information-motivation-behavioral skills model of AIDS-preventive behavior with gay men and heterosexual university students. Health Psychol. 1994;13(3):238–50.

33. Lin W. Agnostic notes on regression adjustments to experimental data: Reexamining Freedman’s critique. Ann Appl Stat. 2013 Mar;7(1):295–318.

34. MacKinnon JG, White H. Some heteroskedasticity-consistent covariance matrix estimators with improved finite sample properties. J Econom. 1985 Sep 1;29(3):305–25.

35. Begg C, Cho M, Eastwood S, Horton R, Moher D, Olkin I, et al. Improving the Quality of Reporting of Randomized Controlled Trials: The CONSORT Statement. JAMA. 1996 Aug 28;276(8):637–9.

36. Li L, Lin C, Guan J. Using standardized patients to evaluate hospital-based intervention outcomes. Int J Epidemiol. 2014 Jun 1;43(3):897–903.

37. Li L, Wu Z, Wu S, Zhao Y, Jia M, Yan Z. HIV-related stigma in health care settings: a survey of service providers in China. AIDS Patient Care STDs. 2007 Oct;21(10):753–62.

38. Li L, Liang L, Lin C, Wu Z, Wen Y. Individual attitudes and perceived social norms: Reports on HIV/AIDS-related stigma among service providers in China: International Journal of Psychology: Vol 44, No 6. Int J Psychol [Internet]. 2009 [cited 2023 Jun 6];44(6). Available from: https://www.tandfonline.com/doi/abs/10.1080/00207590802644774

39. Cai G, Moji K, Honda S, Wu X, Zhang K. Inequality and Unwillingness to Care for People Living with HIV/AIDS: A Survey of Medical Professionals in Southeast China. AIDS Patient Care STDs [Internet]. 2007 [cited 2023 Jun 6];21(8). Available from: https://www.liebertpub.com/doi/epdf/10.1089/apc.2006.0162

40. Wasserman D. Is Racial Profiling More Benign in Medicine Than Law Enforcement? J Ethics. 2011;15(1–2):119–29.

41. Burgess DJ, Warren J, Phelan S, Dovidio J, van Ryn M. Stereotype Threat and Health Disparities: What Medical Educators and Future Physicians Need to Know. J Gen Intern Med. 2010;25(S2):169–77.

42. Betancourt JR, Green AR, Emilio Carrillo J, Park ER. Cultural competence and health care disparities: Key perspectives and trends. Health Aff (Millwood). 2005;24(2):499–505.

43. Harper GW. Sex isn’t that simple: culture and context in HIV prevention interventions for gay and bisexual male adolescents. Am Psychol. 2007;62(8):803–19.

44. Rao D, Elshafei A, Nguyen M, Hatzenbuehler ML, Frey S, Go VF. A systematic review of multi-level stigma interventions: State of the science and future directions. BMC Med. 2019;17(1):1–11.

45. Nyblade L, Mbuya-Brown RJ, Ezekiel MJ, Addo NA, Sabasaba AN, Atuahene K, et al. A total facility approach to reducing HIV stigma in health facilities: implementation process and lessons learned. AIDS Lond Engl. 2020;34(October 2019):S93–102.

46. Ayala G, Sprague L, Leigh-Ann van der Merwe L, Thomas RM, Chang J, Arreola S, et al. Peer-And community-led responses to HIV: A scoping review. PLoS ONE. 2021;16(12 December).

