## Supplemental files for "An Incognito Standardized Patient Approach for Measuring and Reducing Intersectional Healthcare Stigma"

Table S1. Details on the measures for each of the three healthcare quality domains.

| Domains of care | Type | Details | Notes |
|---|---|---|---|
| Syphilis testing | Binary | Whether or not syphilis testing was offered | B section, question 17. |
| Diagnostic effort | Continuous | - 19 indicators combined into a single score including patient history taking items included (e.g. asking patient about fever or past STI diagnoses) and types of physical exam performed (e.g., visual inspection of the patients' hands, torso, or genital area).<br>- All indicators were then combined into a continuous summary index which was weighted using covariance-weighted averages of the included items. Scores were standardized to the sample mean with a mean value of 0 and a standard deviation of 1. | Section B |
| Patient-centered | Continuous | - 19 indicators combined into a single score including items on nonverbal communication (e.g. level of eye contact, how comfortable the provider appeared during the visit), verbal communication (e.g., tone of speech, any mention of discriminatory attitudes towards gay or HIV positive people), and shared decision making using CollaboRATE, a patient-reported scale that has been validated in Chinese language settings (18).<br>- Items for the patient centered care score were also combined into a continuous summary index using the same approach as for clinical management. | Sections C,D,E, and F |

Table S2. The healthcare quality checklist used for data collections during SP-based experimental audits.

| |
|---|
| ***Visit date*** |
| ***Visit time (IN)*** |
| ***Visit time (OUT)*** |
| ***SP name*** |
| ***Facilitator name*** |
| ***Facility name*** |
| ***Provider name*** |

### A. General debrief [ENUMERATOR: AUDIO RECORD]

Please tell me everything that happened. [Allow SP to fully recount encounter while recording]

Can you talk about your experience of the visit as it relates to privacy?

Great, now we will move forward to fill out the checklist together. [Continue to record]

| **B. Adherence to Clinical Guidelines** | | | |
|---|---|---|---|
| **Consultation**: did the doctor ask questions related to any of the following? | | | |
| 1. | Symptoms | | |
| | *1.1 Chancre in genital area* | | 1=Yes 2=No |
| | *1.2 Enlarged lymph nodes* | | 1=Yes 2=No |
| | *1.3 Fever* | | 1=Yes 2=No |
| | *1.4 Malaise* | | 1=Yes 2=No |
| | *1.5 Hair Loss* | | 1=Yes 2=No |
| | *1.6 Muscle or joint pain* | | 1=Yes 2=No |
| | *1.7 Loss of appetite* | | 1=Yes 2=No |
| | *1.8 Mouth Ulcer* | | 1=Yes 2=No |
| 2. | Other STD Related Symptoms | | |
| | *2.1 Have you ever diagnosed any STD?* | | 1=Yes 2=No |
| | *2.2 Penis: Bleeding* | | 1=Yes 2=No |
| | *2.3 Penis: Secretion* | | 1=Yes 2=No |
| | *2.4 Penis: Blister* | | 1=Yes 2=No |
| | *2.5 Rectum: Growth/Tumor* | | 1=Yes 2=No |
| | *2.6 Rectum: Swelling/redness* | | 1=Yes 2=No |
| | *2.7 Rectum: Secretion* | | 1=Yes 2=No |
| 3. | HIV-related | | |
| | *3.1 HIV infection status* | | 1=Yes 2=No |
| | *3.2 If positive: Most recent CD4 count* | | 1=Yes 2=No |
| | *3.3 If positive: Current treatment status* | | 1=Yes 2=No |
| 4. | Sexual behaviors | | |
| | *4.1 Number of partners* | | 1=Yes 2=No |
| | *4.2 Condom use* | | 1=Yes 2=No |
| | *4.3 Commercial sex* | | 1=Yes 2=No |
| | *4.4 HIV status of sexual partners* | | 1=Yes 2=No |
| | *4.5.Same sex behaviors* | | 1=Yes 2=No |
| 5. | Alcohol use | | 1=Yes 2=No |
| 6. | Recreational drug use (e.g. Poppers/Rush, Ecstasy) | | 1=Yes 2=No |
| **Exam**: Did provider perform (or offer to perform) physical exams of: | | | |
| 7. | Hands | | 1=Yes 2=No |
| 8. | Torso | | 1=Yes 2=No |

| | | | |
|---|---|---|---|
| 9. | Lymph nodes | | 1=Yes 2=No |
| 10. | Genital area (Offer only, SPs should refuse) | | 1=Yes 2=No |
| **Counseling**: Did the provider initiate counselling on any of following topics? | | | |
| 11. | Advice on reducing partner numbers | | 1=Yes 2=No |
| 12. | Advice on condom use | | 1=Yes 2=No |
| 13. | Referral of partners for testing (HIV or other STD) | | 1=Yes 2=No |
| 14. | Abstain from sex for a short period or before test result comes up | | 1=Yes 2=No |
| **Procedures**: Did provider make diagnosis, order tests, or prescribe drugs? | | | |
| 14. | Was diagnosis given? | | 1=Yes 2=No |
| 15. | Were drugs prescribed? | | 1=Yes 2=No |
| 16. | Syphilis test ordered | | 1=Yes 2=No |
| 17. | HIV test ordered | | 1=Yes 2=No |
| 18. | Other STD test ordered | | 1=Yes 2=No |
| **C. Clinic Experience** | | | |
| 1. | How long was your interaction with the doctor? | | minutes |
| 2. | How long did you wait before seeing the provider? | | minutes |
| 3. | Were you refused or denied care? *[If NO, skip to Question 5]* | | 1=Yes 2=No |
| 4. | If denied care, which reason(s) were given? | *Check all that apply* | |
| | *4.1 Facility does not provide services needed* | | |
| | *4.2 Facility not prepared to treat patients with HIV* | | |
| | *4.3 Other facility better equipped to treat HIV infected patients* | | |
| | *4.4 No reason given* | | |
| | *4.5 Other* | | |
| | *4.5.1: Specify:* | | |
| 5. | Did the examination take place in a room with a door? | | 1=Yes 2=No |
| | If Yes, was the door open or closed? | | 1=open<br>2=closed |
| 6. | Were other people within earshot during the visit? | | 1=Yes 2=No |
| 7. | Was the doctor interrupted by someone else for any reason during the visit? | | 1=Yes 2=No |
| **D. Nonverbal Communication** | | | |
| 1. | How would you describe the level eye contact you had with the provider? | | 1=Less<br>2=Somewhat less<br>3=Somewhat lot<br>4=Lot |
| 2. | Did the provider seem comfortable during the interaction? | | 1=Very uncomfortable<br>2=Uncomfortable<br>3=Comfortable<br>4=Very comfortable |
| 3. | Did the provider make you feel like he/she was genuinely concerned for your health problem? | | 1=No<br>2=Somewhat not concern<br>3=Somewhat concern<br>4=Concern |
| 4. | Did the provider address your concerns about transmitting an STD to your boyfriend/girlfriend? | | 1=Yes<br>2=No |

| | | | |
|---|---|---|---|
| 5. | How would you describe the provider's body posture? (includes cues such as crossed arms, leaning in or out, fidgeting, etc.) | | 1=Closed<br>2=Somewhat closed<br>3=Somewhat open<br>4=Open |
| 6. | How would you describe the provider's physical presentation? | | 1=Tense<br>2=Somewhat tense<br>3=Somewhat relaxed<br>4=Relaxed |
| 7. | How close did the provider stand/sit to you? (proxemics) | | 1=Somewhat near<br>2=Near<br>3=Somewhat far<br>4=Far |
| **E. Verbal Communication** | | | |
| 1. | How would you describe the provider's tone of speech?* | | 1=Positive<br>2=Negative<br>3=Neutral |
| 2. | To your knowledge, did the provider disclose any patient information to others (other clinic staff, other patients) without SP consent? | | 1=Yes 2=No |
| 3. | Beliefs | | |
| 3.1 | [For MSM case scenarios]: Did the provider say or do anything that implied any of the following beliefs? | | |
| | *3.1.a All or most gay men have HIV* | | 1=Yes 2=No |
| | *3.1.b Gay men have HIV because they are gay* | | 1=Yes 2=No |
| | *3.1.c Same sex behaviors are immoral (e.g. inappropriate, strange, repulsive, unnatural)* | | 1=Yes 2=No |
| | *3.1.d Same sex behaviors are unhealthy* | | 1=Yes 2=No |
| | *3.1.e Same sex behaviors should be banned or made illegal* | | 1=Yes 2=No |
| 3.2 | [For HIV positive case scenarios]: Did the provider say or do anything that implied any of the following beliefs? | | |
| | *3.2.a People with HIV got what they deserved* | | 1=Yes 2=No |
| | *3.2.b HIV infection results from inappropriate or immoral behaviors* | | 1=Yes 2=No |
| | *3.2.c People with HIV should not receive medical care in the same place as uninfected patients* | | 1=Yes 2=No |
| | *3.2.d People with HIV should not have sex* | | 1=Yes 2=No |
| | *3.2.e People with HIV should not have children* | | 1=Yes 2=No |
| ** Positive tone: warm, supportive, understanding, empathetic; Negative tone: nervous, disgusted, agitated, judgmental* | | | |

**F. Shared Decision Making**

| | | |
|---|---|---|
| 1 . | How much effort do you think the doctor make during helping you solve your problem? | 0-9 （0= no effort, 9=give his/her all effort） |
| 2. | How much effort do you think the doctor make during listening to your most concern issue? | 0-9 （0= no effort, 9=give his/her all effort） |
| 3. | When making next step decision, how much effort do you think the doctor provide to consider your most concern issue? | 0-9 （0= no effort, 9=give his/her all effort） |
